# Therapeutic Management of COVID-19 Patients: A systematic review

**DOI:** 10.1101/2020.04.02.20051029

**Authors:** Mansour Tobaiqy, Mohammed Qashqary, Shrooq Al-Dahery, Alaa Mujallad, Almonther Abdullah Hershan, Mohammad Azhar Kamal, Nawal Helmi

## Abstract

**Background:** SARS-CoV-2 is the cause of the COVID-19 that has been declared a global pandemic by the WHO in 2020. The COVID-19 treatment guidelines vary in each country, and yet there is no approved therapeutic for COVID-19.

**Aims of the study:** this review aimed to report any evidence of therapeutics used for the management of COVID-19 patients in clinical practice since the emergence of the virus.

**Methods:** A systematic review protocol was developed based on PRISMA Statement. Articles for review were selected from electronic databases (Embase, Medline and Google Scholar). Readily accessible peer-reviewed full articles in English published from December 1 ^st^, 2019 to March 26 ^th^, 2020 were included. The search terms included combinations of: COVID, SARS-COV-2, glucocorticoids, convalescent plasma, antiviral, antibacterial. There were no restrictions on the type of study design eligible for inclusion.

**Results:** As of March 26, 2020, of the initial manuscripts identified (n=449) articles. Forty-one studies were included, of which clinical trials (n=3), (case reports n=7), case series (n=10), retrospective (n=11) and prospective (n=10) observational studies. Thirty-six studies were conducted in China (88%).

The most common mentioned and reported medicine in this systematic review was corticosteroids (n=25), followed by Lopinavir (n=21) and oseltamivir (n=16).

**Conclusions:** This is the first systematic review up to date related to the therapeutics used in COVID-19 patients. Only forty-one research articles on COVID-19 and therapeutics were found eligible to be included, most conducted in China, corticosteroid therapy was found to be the most used medicine in these studies.

## Introduction

Severe acute respiratory syndrome coronavirus SARS-CoV-2 is the cause of the coronavirus disease 2019 (COVID-19) that has been declared a global pandemic by the World Health Organization (WHO) in 2020. SARS-CoV-2 was discovered in December of 2019, in Wuhan City in the capital of Hubei province in China. The origin of the virus is unknown, but all the newly diagnosed cases are linked to the Huanan Seafood Wholesale Market where people can buy illegal wild animals, such as bats ^(1)^. The pathogen of SARS-CoV-2 has the same phylogenetic similarity to SARS-CoV and MERS-CoV. The virus was identified as a novel enveloped RNA betacoronavirus that has been named as Severe Acute Respiratory Syndrome Coronavirus 2 (SARS-CoV-2) ^(2)^.

One of the characteristics of COVID-19 is it is highly contagious; many countries were affected, including China and 164 other countries within less than three months. Despite China reaching 81,151 confirmed cases with 3,242 deaths, the country reported only one new domestic case as of March 18^th^, 2020; as of that date, the total worldwide confirmed cases are 193,475 with 7,864 deaths (WHO). Although protective measures have been implemented in China, such as isolation from confirmed and suspected cases to reduce the spread of the virus, the need for effective treatment is imperative to stop the outbreak of COVID-19 ^(1)^.

Since the outbreak, researchers have released many agents that could have a potential efficacy against COVID-19. Not only did western medicine start testing some agents in clinical studies to observe the effectiveness of the medications, but they are also engaging natural products and Chinese medicines. Different types of antivirals were included in the latest guidelines from the National Health Commission (NHC) including interferon, lopinavir/ritonavirus, chloroquine phosphate, ribavirin, and arbidol ^(3)^. Angiotention receptor blockers, such as losartan, are another suggestion to treat COVID-19, which would likely be resistant to any new mutation of coronavirus ^(4)^.

The COVID-19 treatment guidelines vary in each country. The WHO guidelines are very general to manage only the symptoms and advised to be cautious with pediatrics, pregnant, and patients with underlying conditions. There is no approved treatment for COVID 19, the care advised is to give according to each patient’s need; Such as antipyretics for fever, and oxygen therapy for patients with respiratory distress. Moreover, the WHO recommendations for the severe cases are to give empiric antimicrobial and implement mechanical ventilation depending on the patient’s situation. Some of the Asian guidelines were not easy to interpret because they are not yet translated to English, such as the Japanese guidelines. The treatment protocols across the countries are nearly similar. They are using hydroxychloroquine, chloroquine phosphate, remedesivir, and lopinavir/ritonavir ^(5-7)^. There are slight differences between some countries treatment guidelines which will be represented in the table 1^(8-11)^.

**Table 1:**
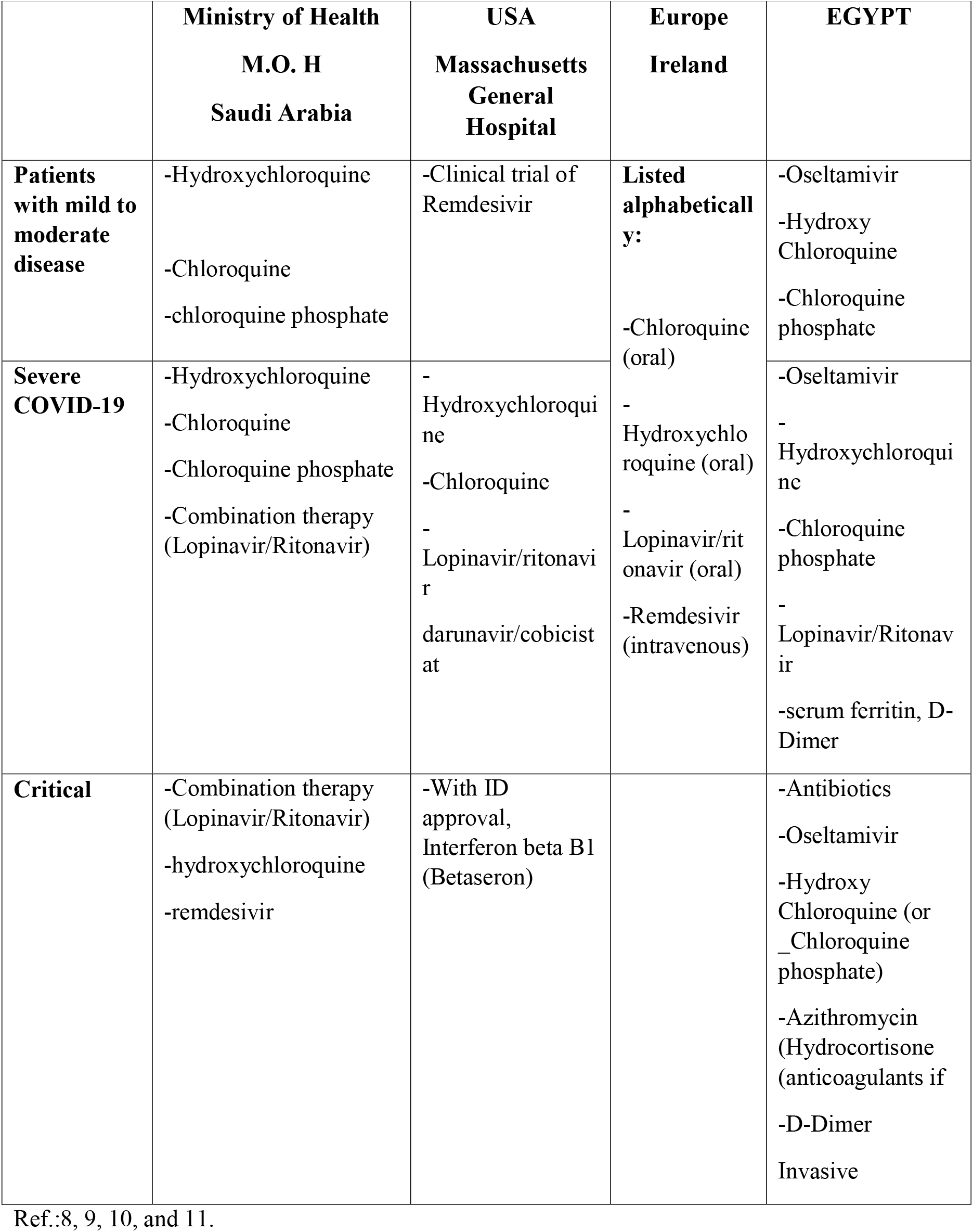
Comparison between the Treatment Guidelines for COVID-19 in Saudi Arabia, USA, Europe, and Egypt

In light of limited and scarce evidence around therapeutics for COVID-19 in the literature, this review aims to retrospectively evaluate the therapeutic management that was given to the COVID-19 patients since the emergence of the virus.

## Methods

A systematic review protocol was developed based on PRISMA-P and the PRISMA Statement. Articles for review were selected from electronic databases (Embase, Medline and Google Scholar). Readily accessible peer-reviewed full articles in English published from December 1^st^, 2019 to March 26^th^, 2020 were included. The search terms included combinations of: COVID-19, SARS-COV-2, glucocorticoids, chloroquine, convalescent plasma, antiviral, antibacterial, oseltamivir, hydroxychloroquine, chloroquine phosphate, monoclonal antibodies. There were no restrictions on the type of study design eligible for inclusion; however, these were likely to be quantitative and RCT articles. The focus of the review was of therapeutics use for the management of COVID-19 patients. Primary outcomes were:

1. the evidence of therapeutics used for the management of COVID-19 patients in clinical practice, since this emergence of the outbreak irrespective of patient characteristics, setting and outcome measures, and to discuss the most common reported medicines in this review.
2. the clinical outcome of the therapeutic treatment (Recovery, Mortality) in COVID-19 patients. Secondary outcomes were adverse events associated with the treatment.

Duplicate articles were removed. Titles were independently screened by both reviewers with abstracts followed by full articles reviewed where any doubt remained. Inclusions and exclusions were recorded following PRISMA guidelines presented in the form of a PRISMA Flow Diagram and detailed reasons recorded for exclusion. Critical appraisal checklists appropriate to each study design were to be applied and checked by a second team member. Any bias or quality issues identified were to be considered prior to a quantitative meta-analysis and meta-narrative, CAPS Appraisal Checklists tools were used for quality assessments. A data extraction tool was designed to capture focus of interest, population, geographical location, methodology, specific mention of therapeutic treatment and adverse events, key findings and further research. Ethical review was not required for this review of existing peer reviewed literature.

## Results

As of March 26, 2020, the initial manuscripts identified (n=449) articles. Inclusions and exclusions are reported following PRISMA guidelines presented in the form of a PRISMA Flow Diagram (Figure 1) with reasons for exclusion recorded (Table 2) as follows: duplicates removed (n=213), records after duplicates removed (n=236), records excluded (n=28) of which n=18 excluded because of the language (n=9 Chinese, n=2 Dutch, n=1 Vietnamese, n=1Spanish, n=1 Italian, n=1 Russian, n=1 Portuguese, n=1 Iranian and n=1 German), ten for other reasons including incomplete and irrelevant articles.

**Table 2.**
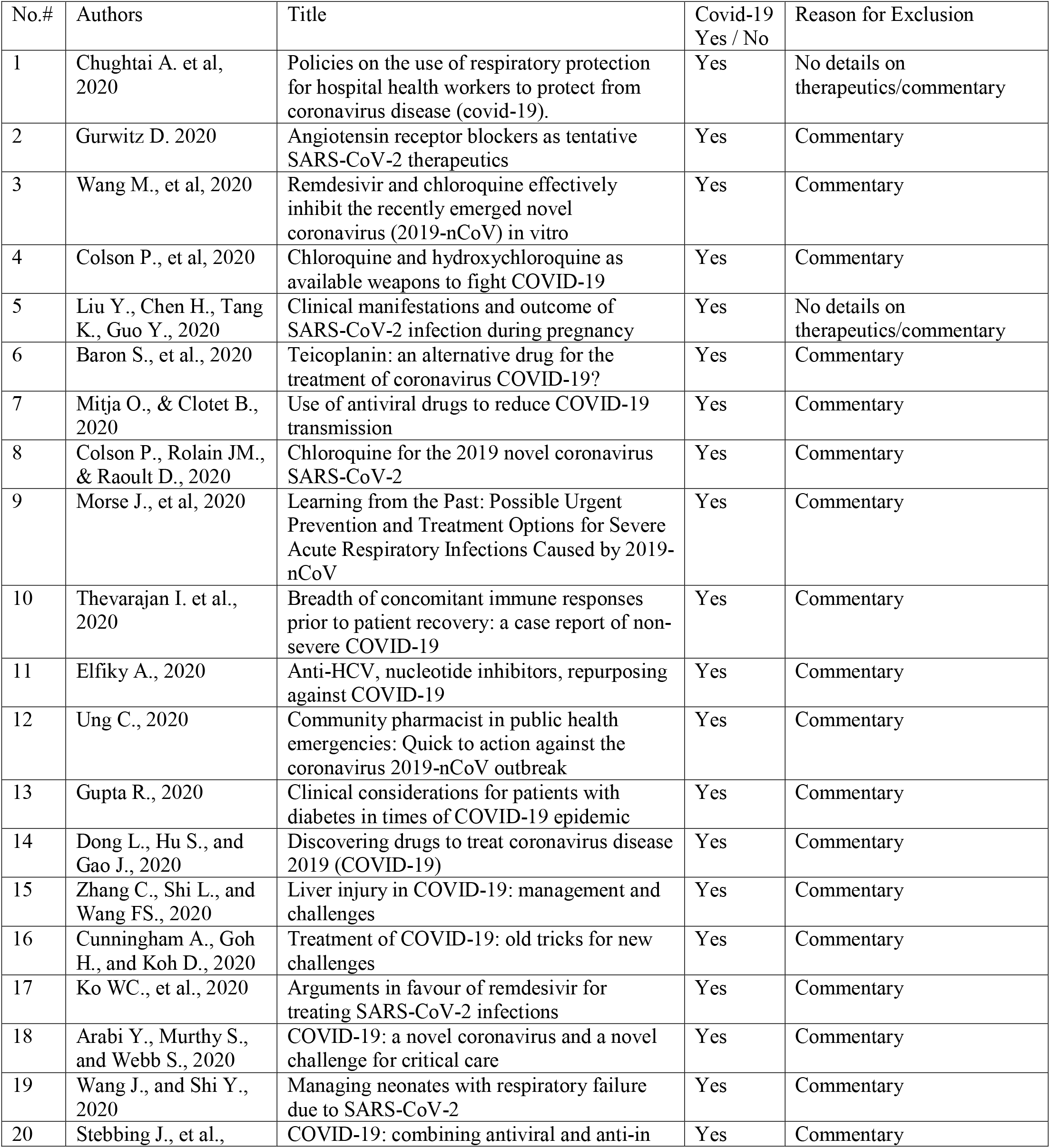
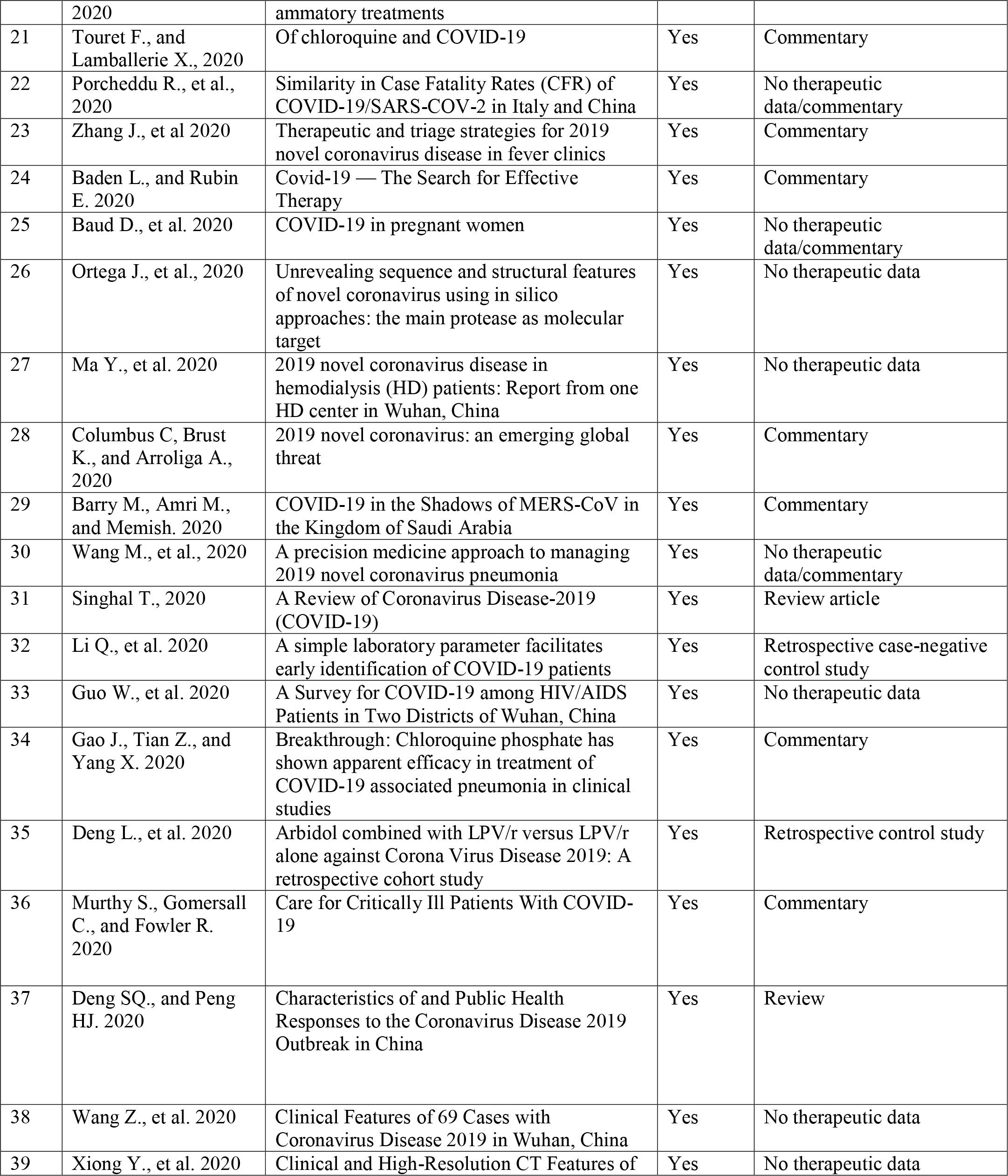
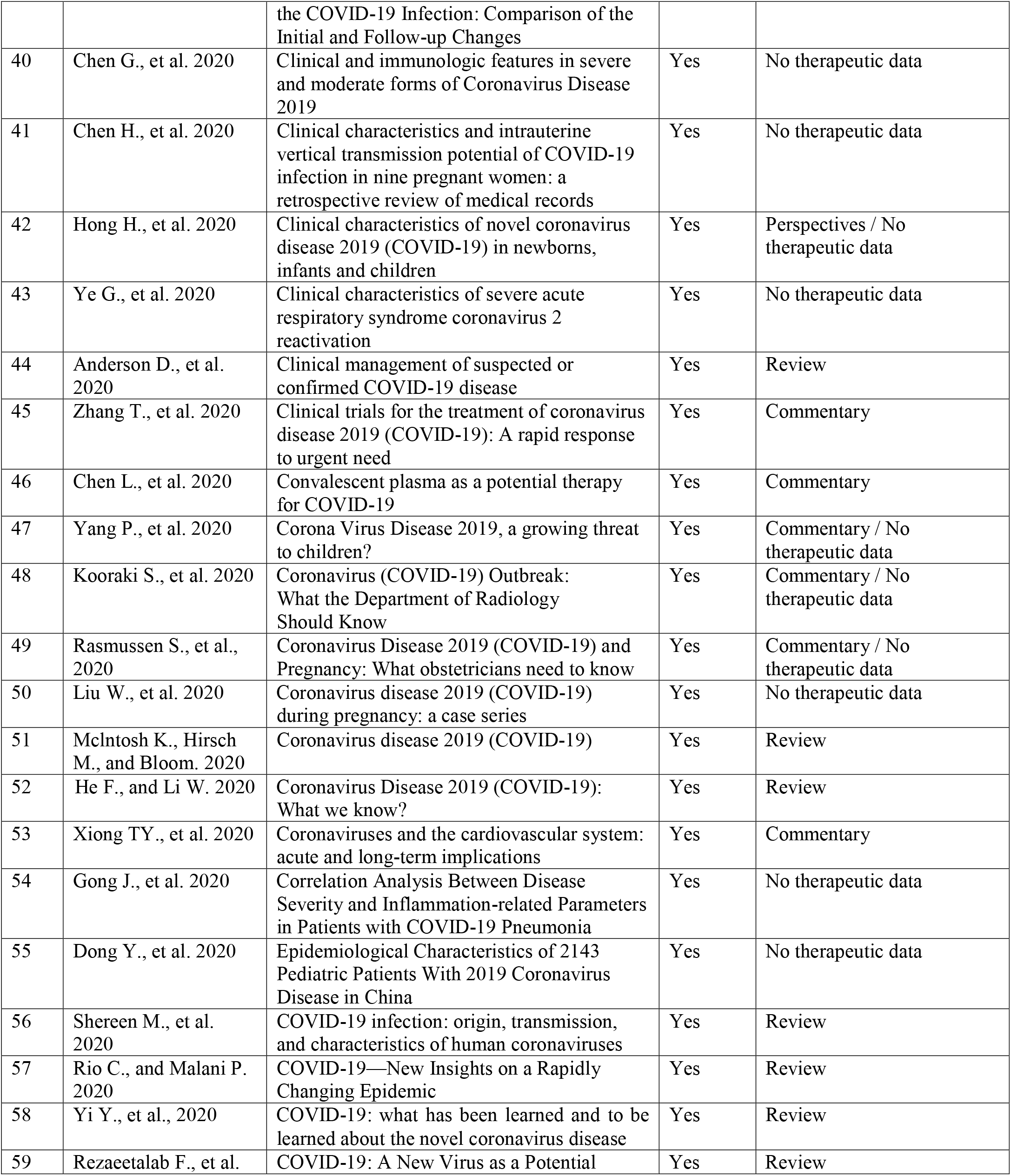
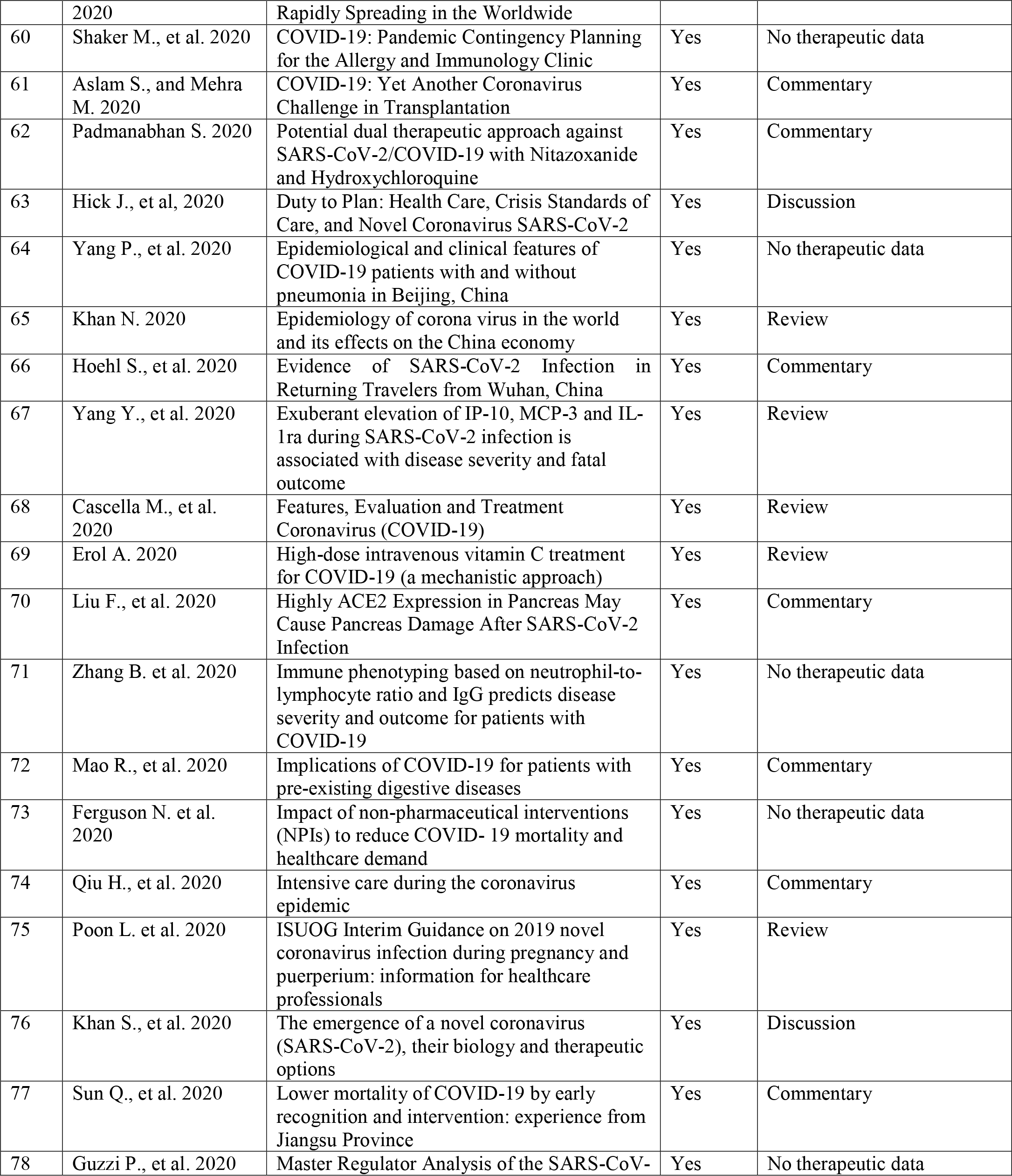
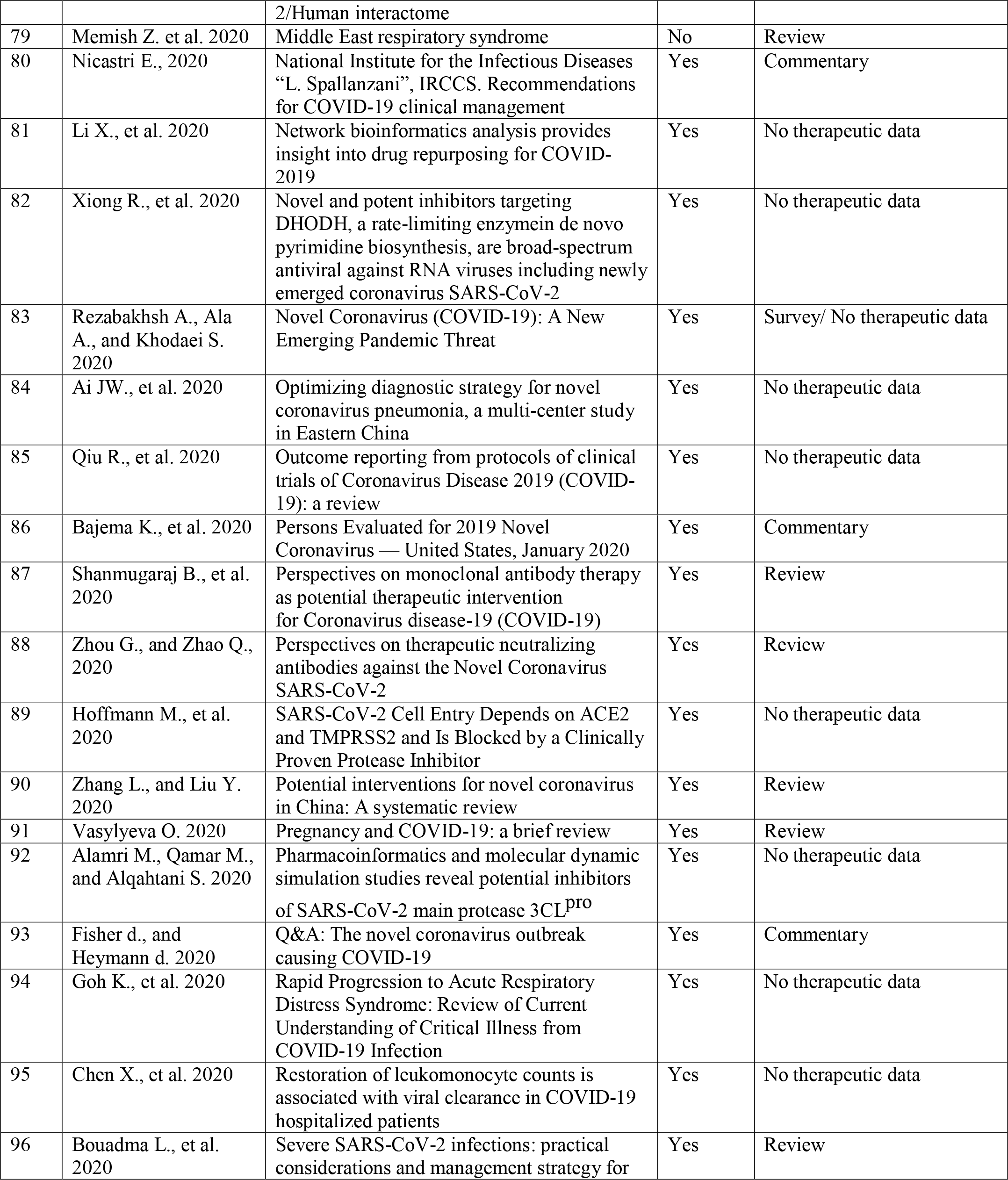
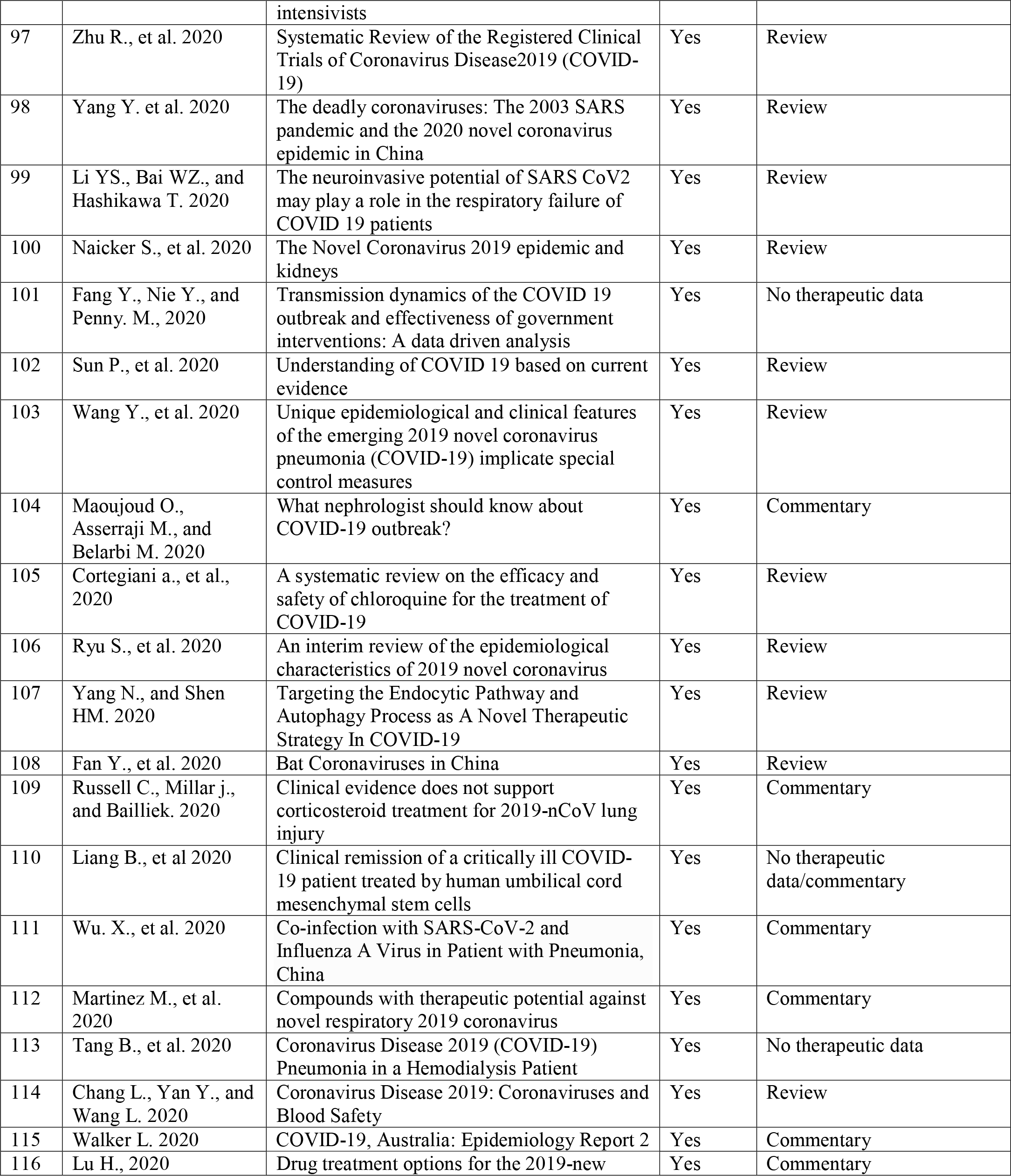
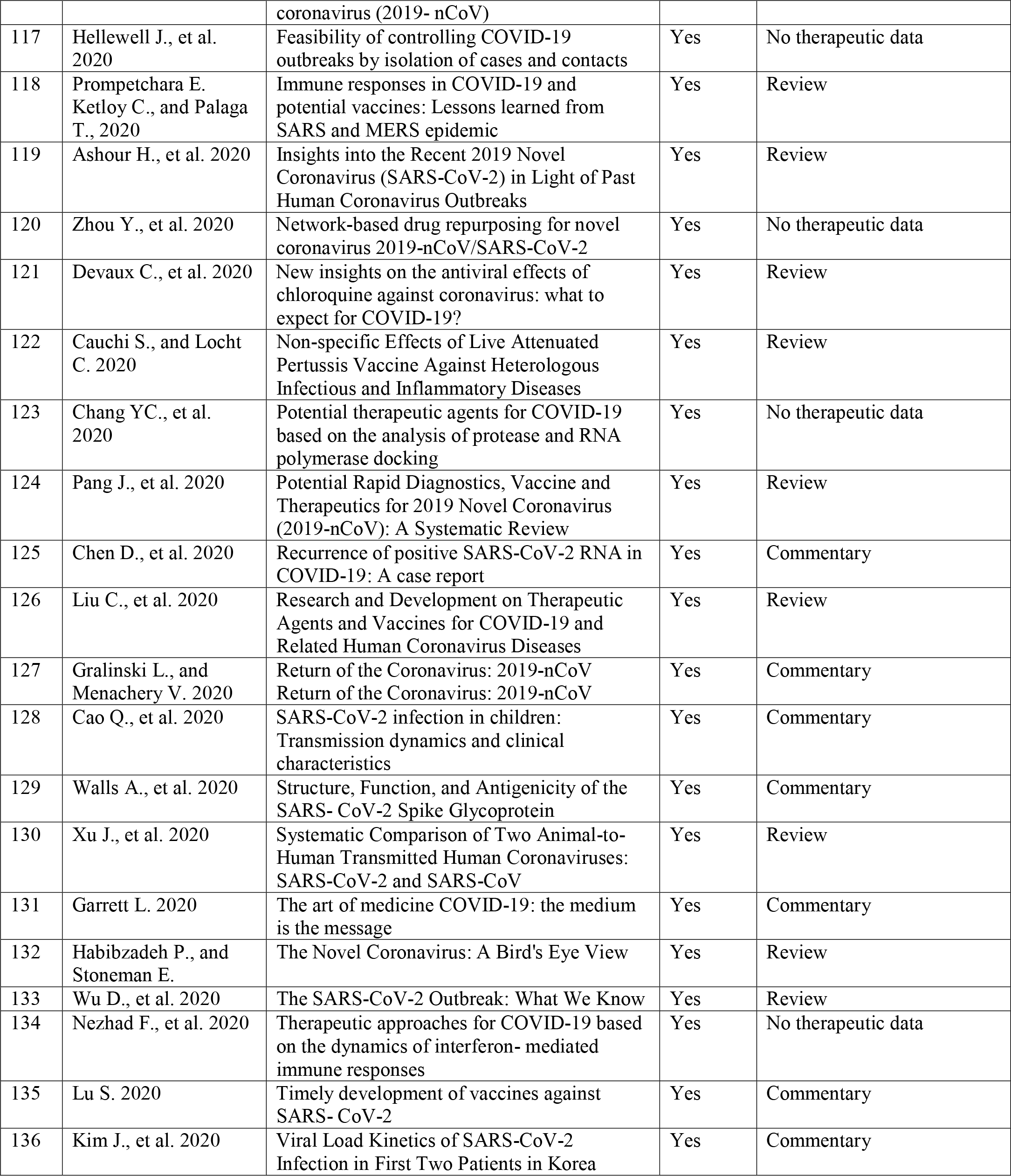
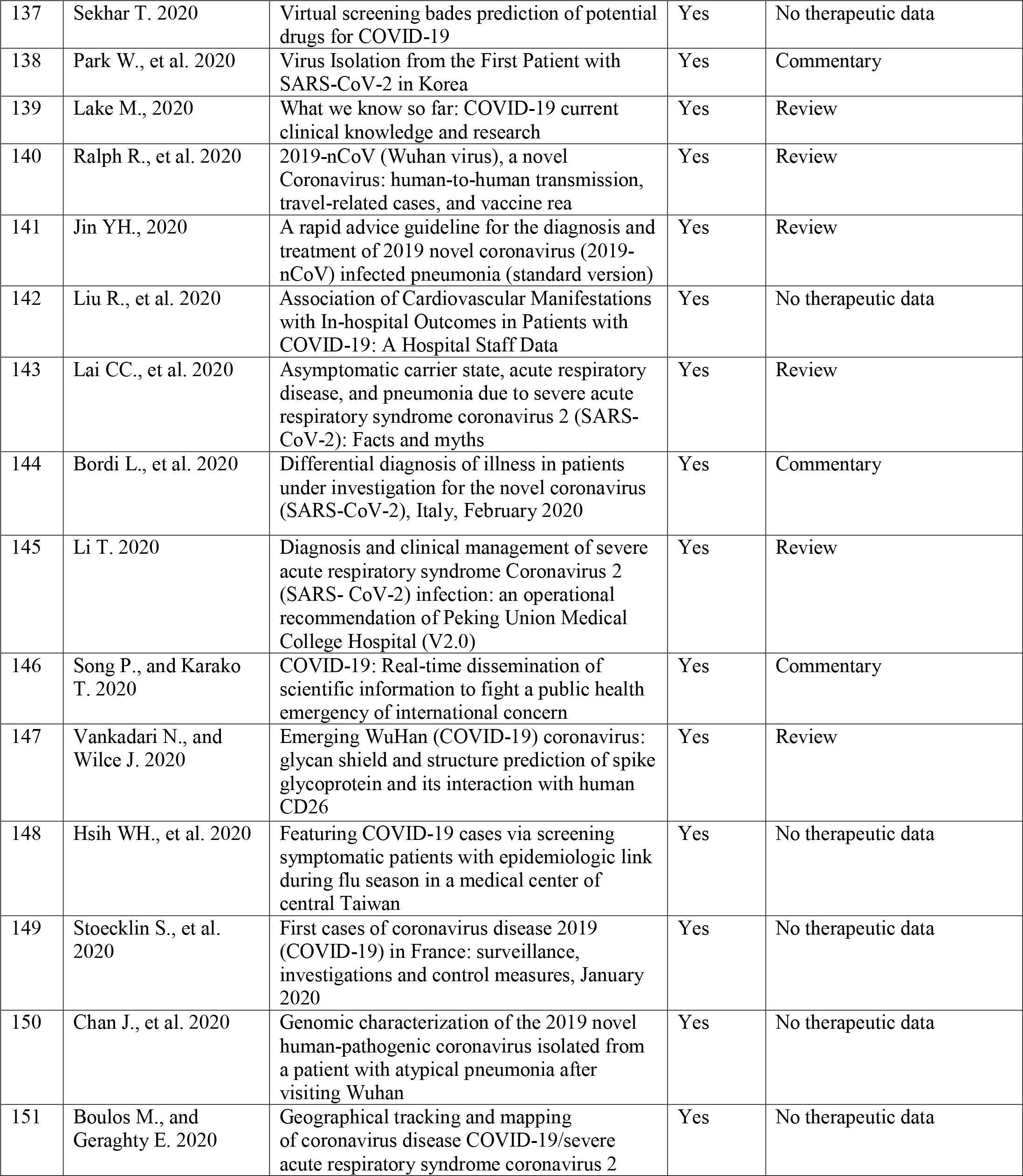
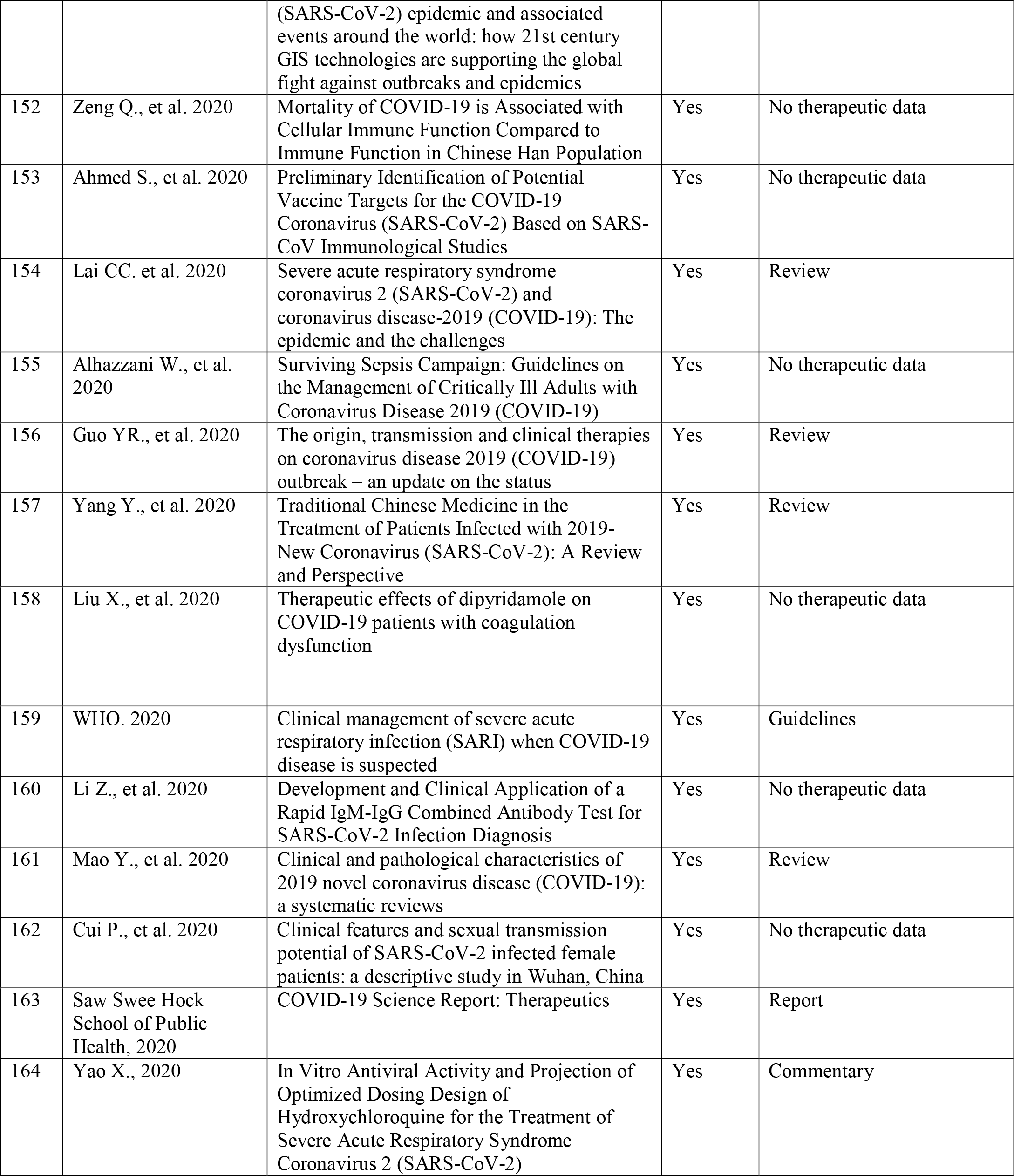
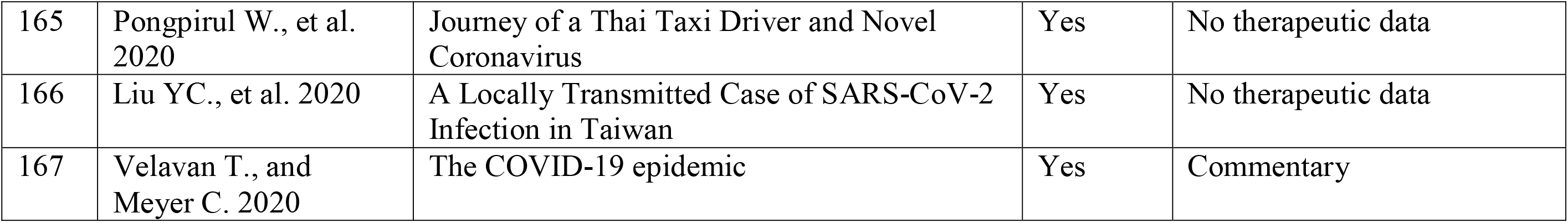
List of excluded papers and reasons for exclusion:

**Figure 1.**
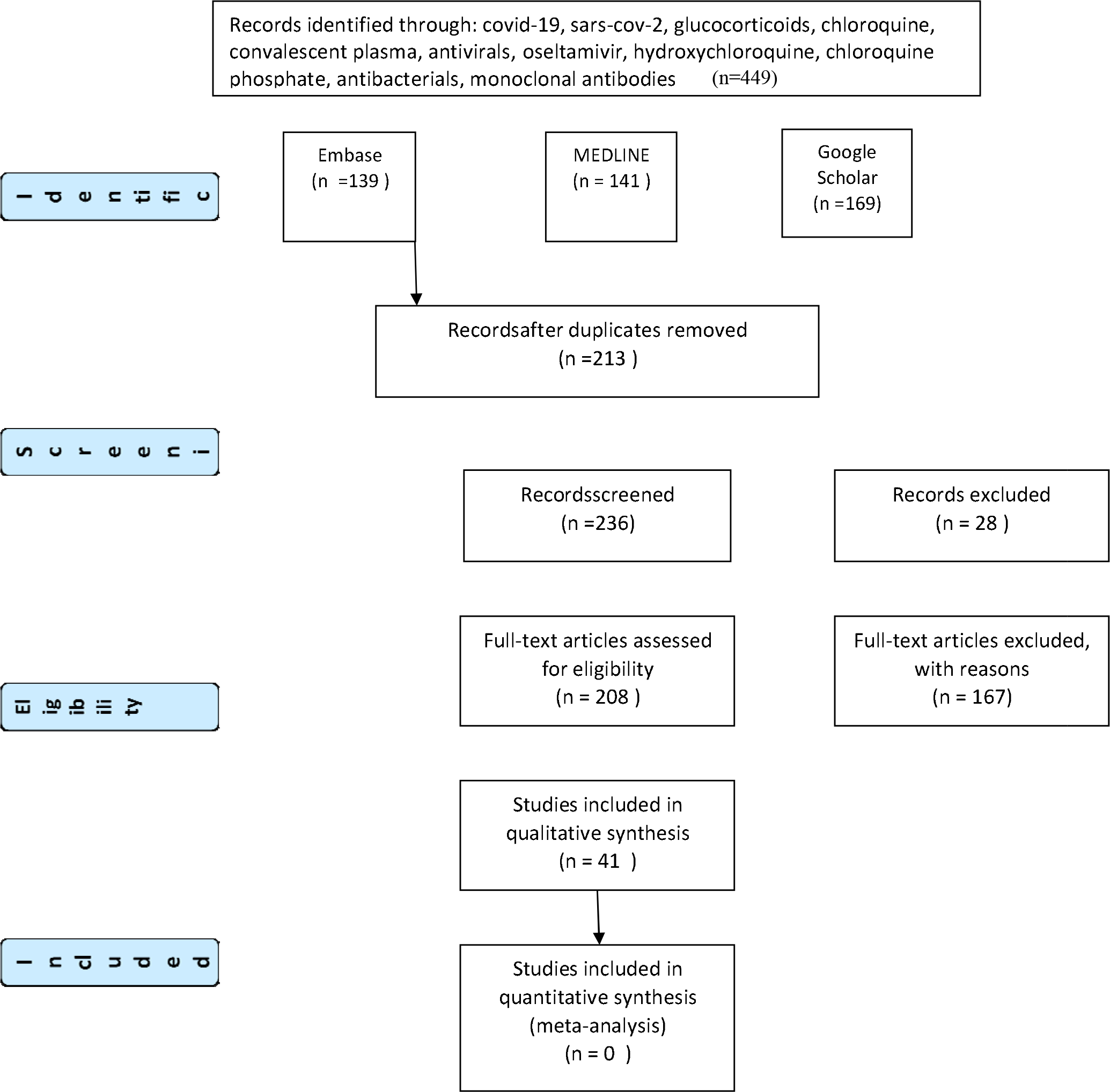
PRISMA Flow Diagram reporting search results.

Consensus on final inclusions (n=41) studies and negotiated without the need for a third reviewer is presented in Table 3.

**Table 3.**
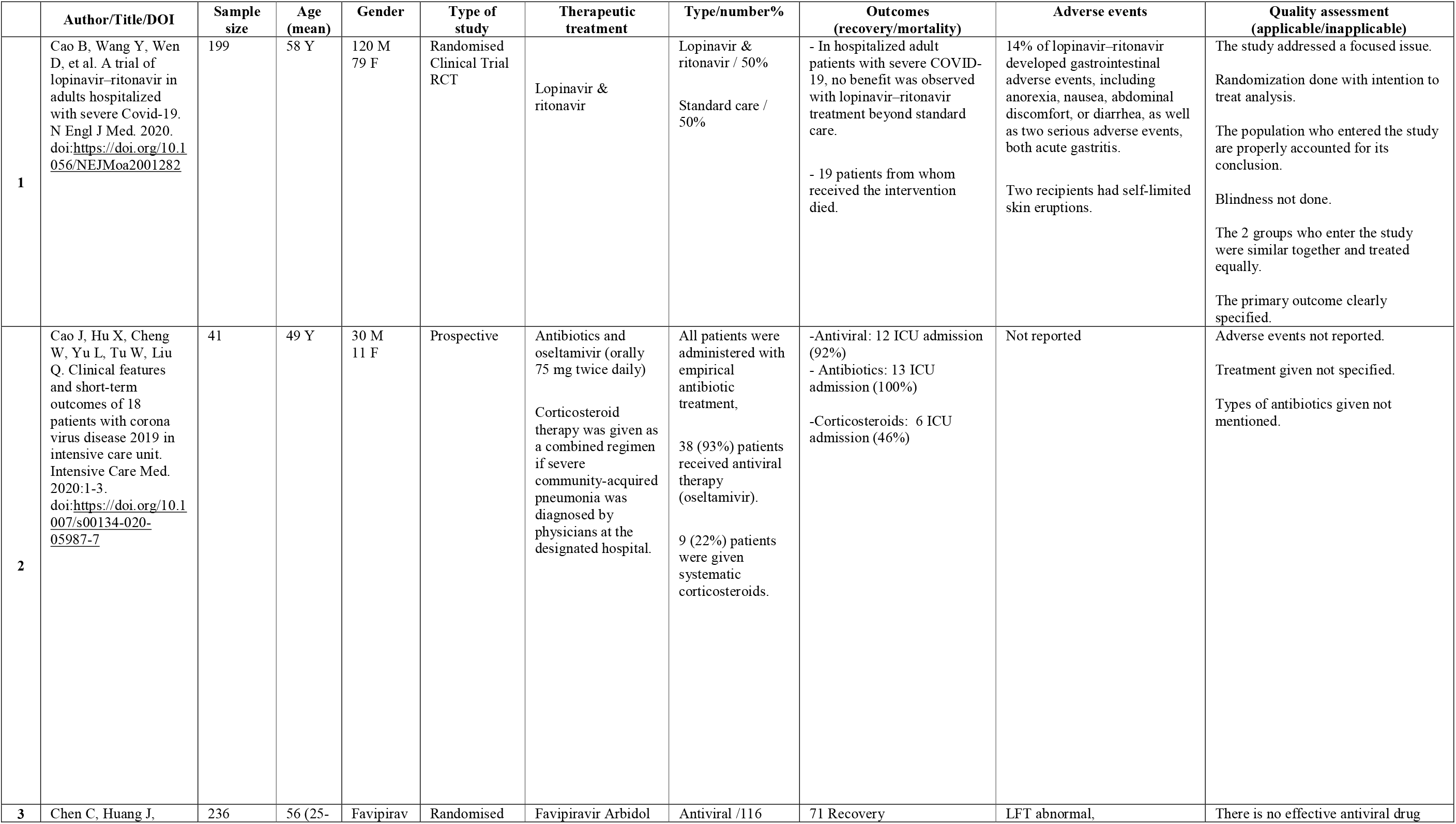
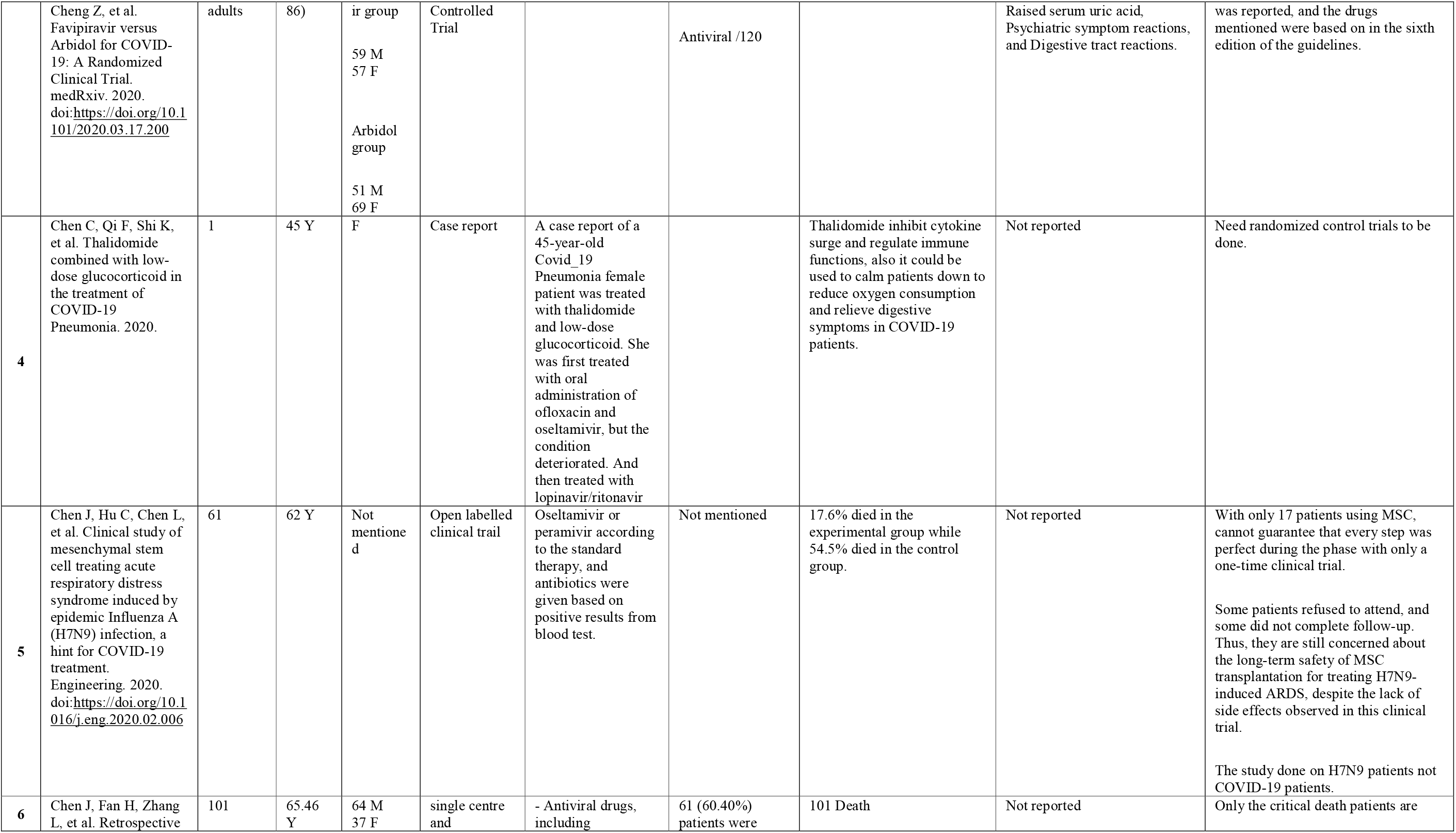
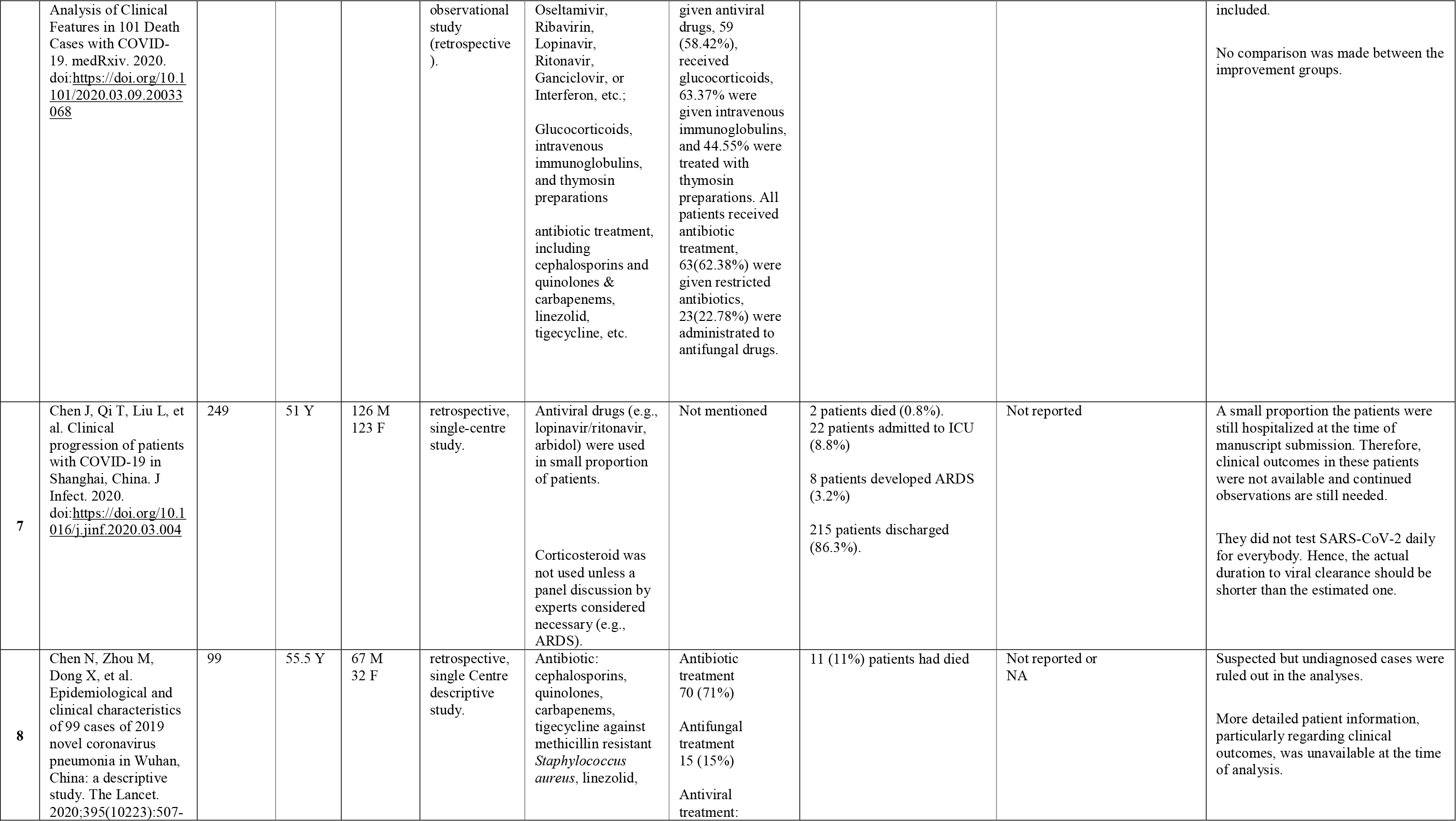
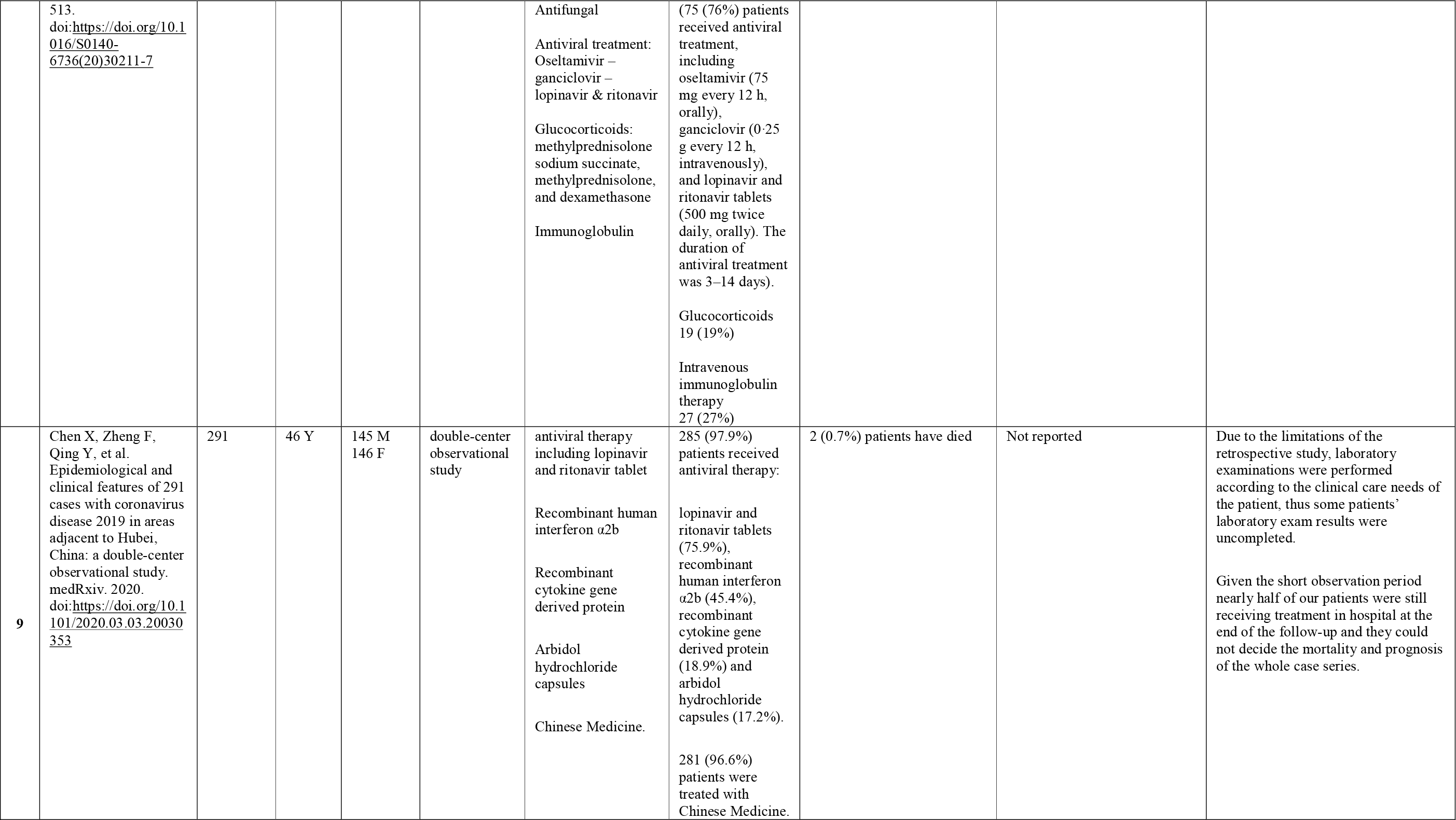
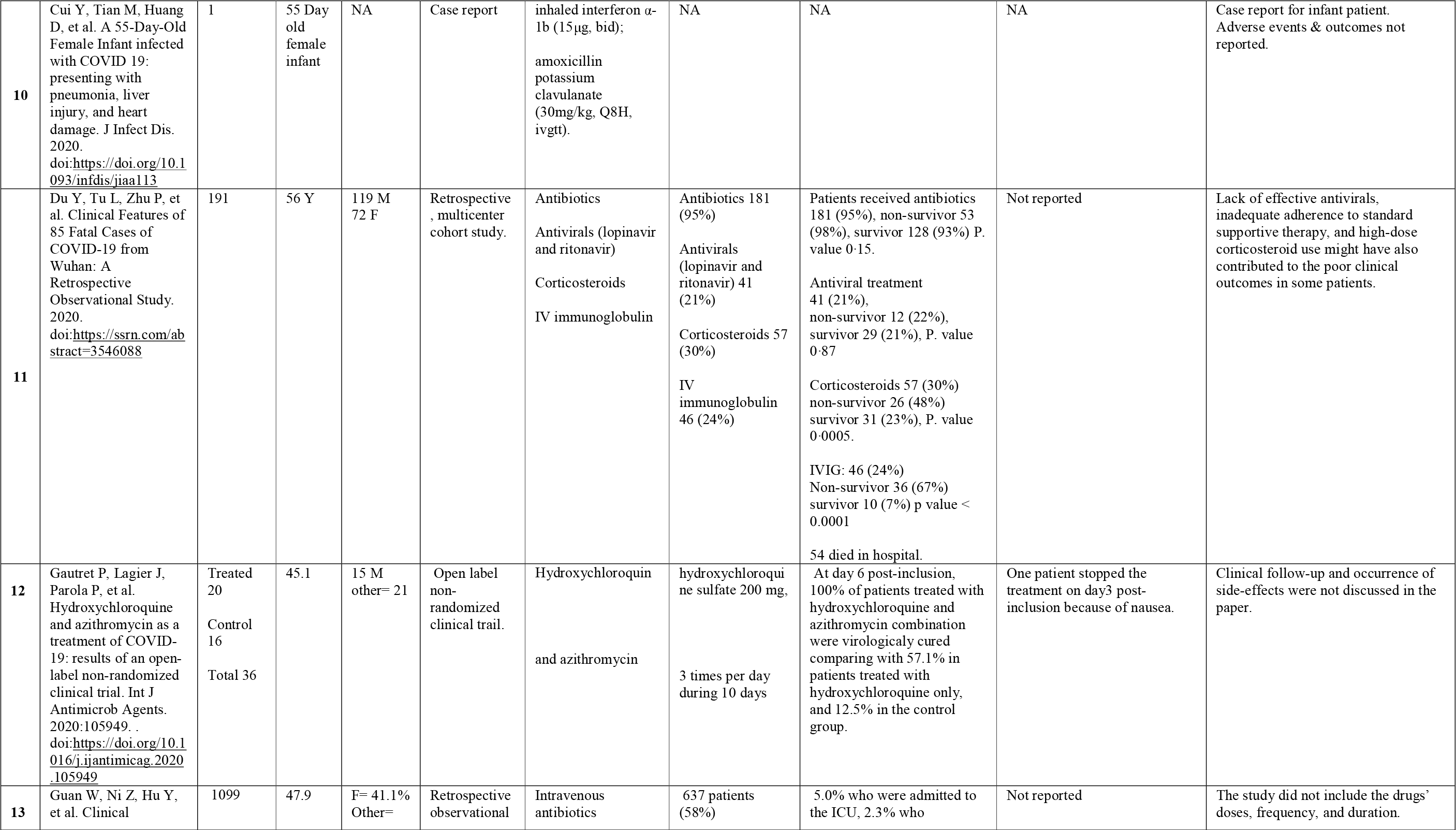
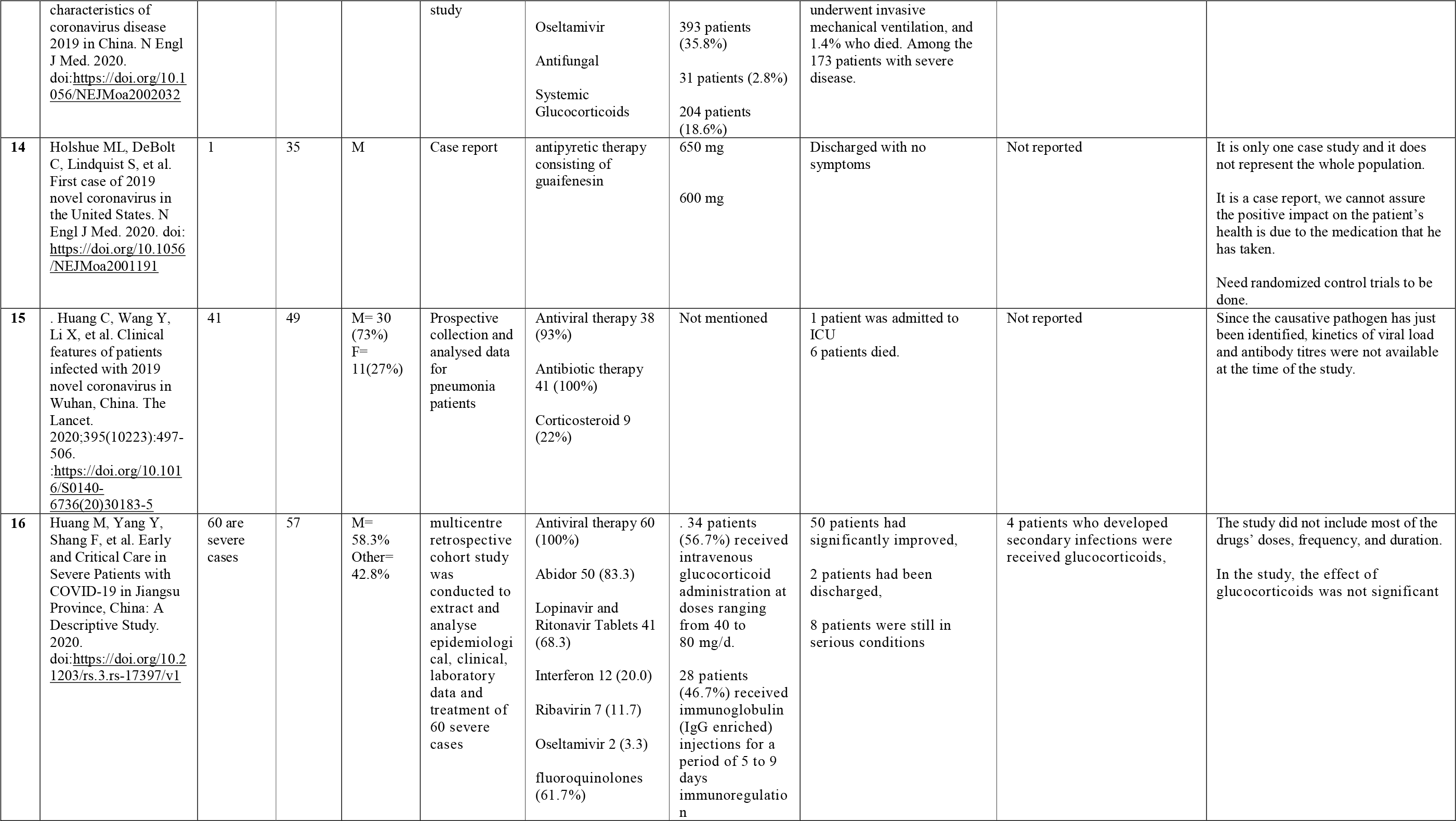
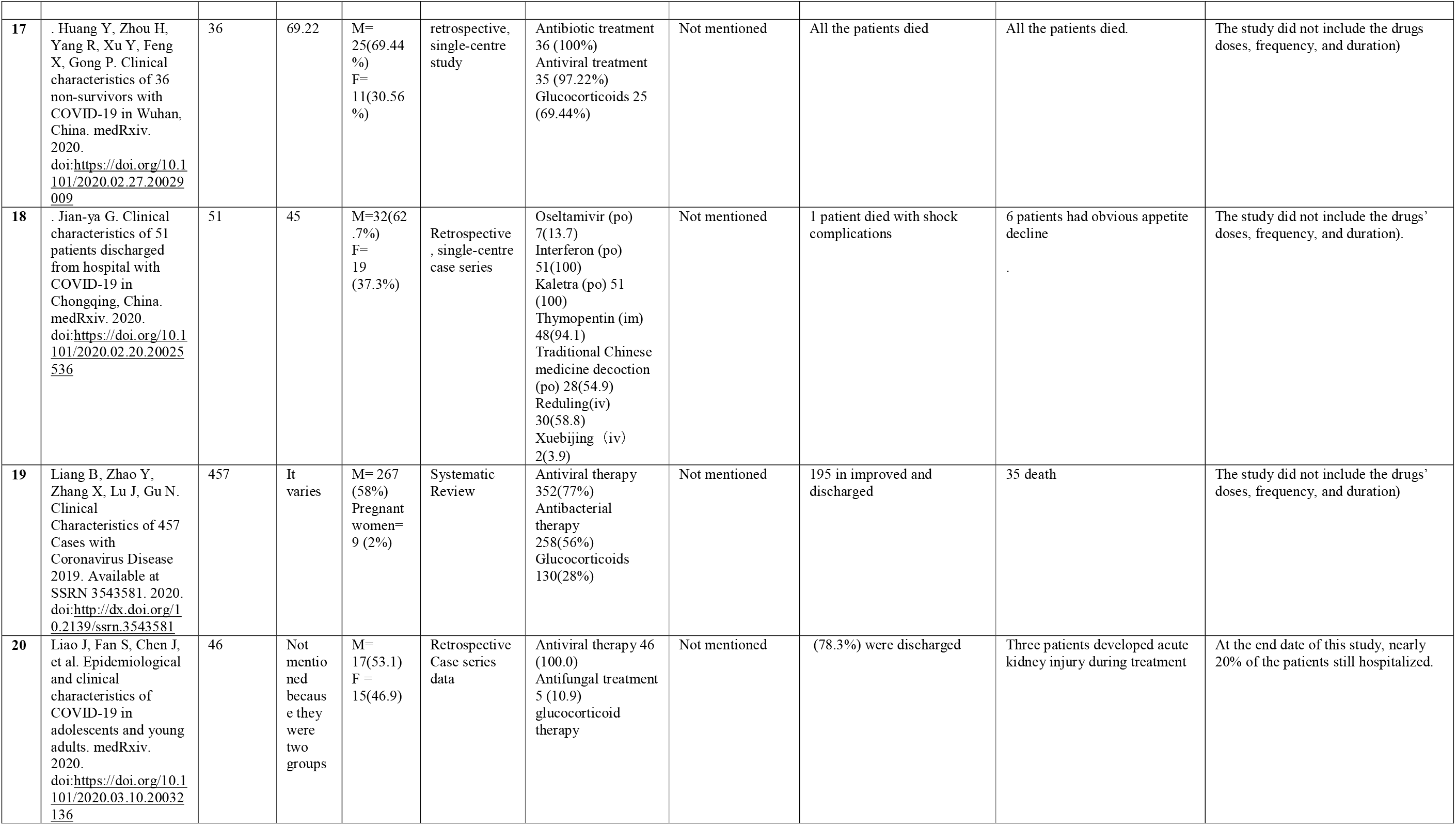
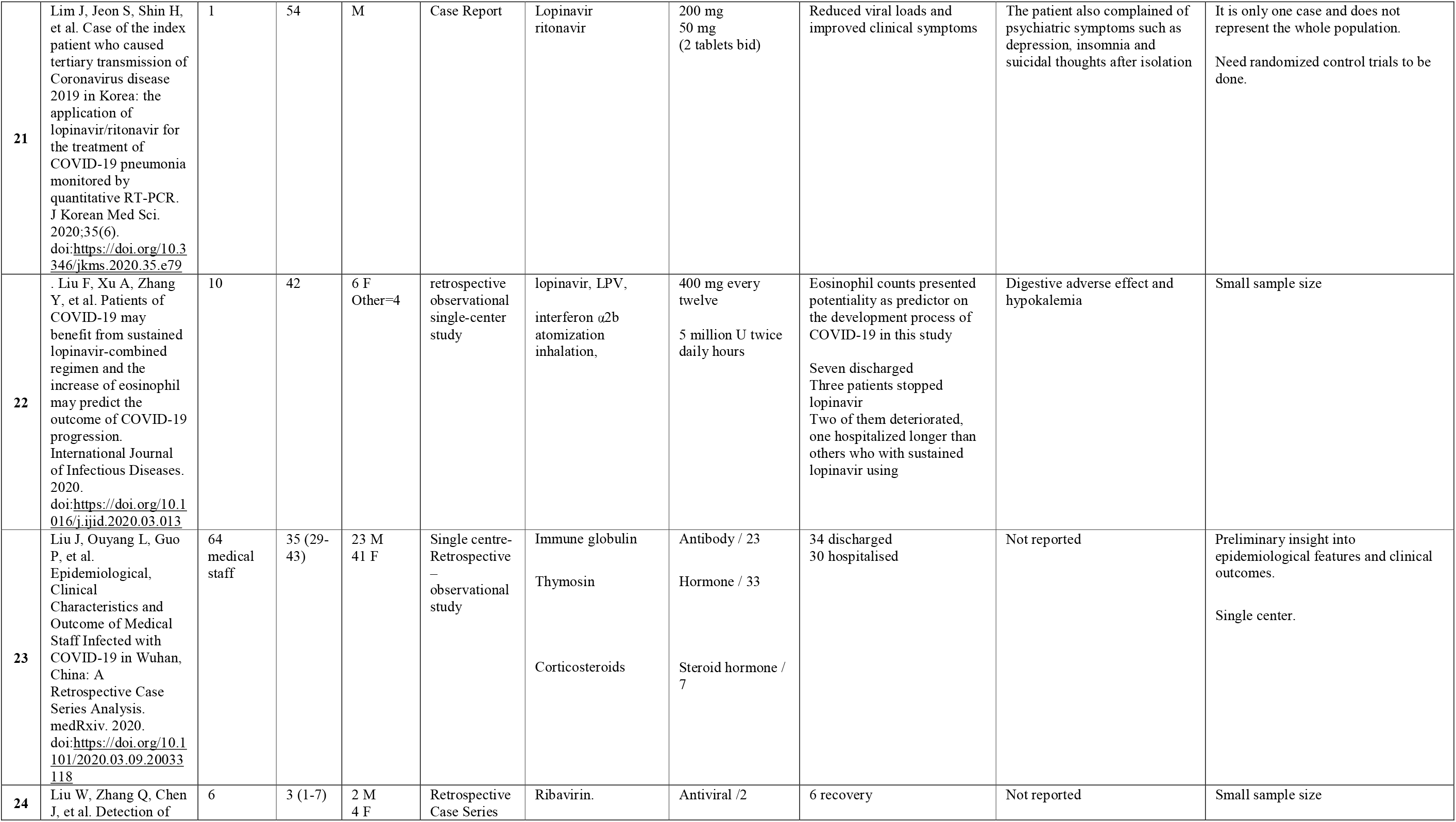
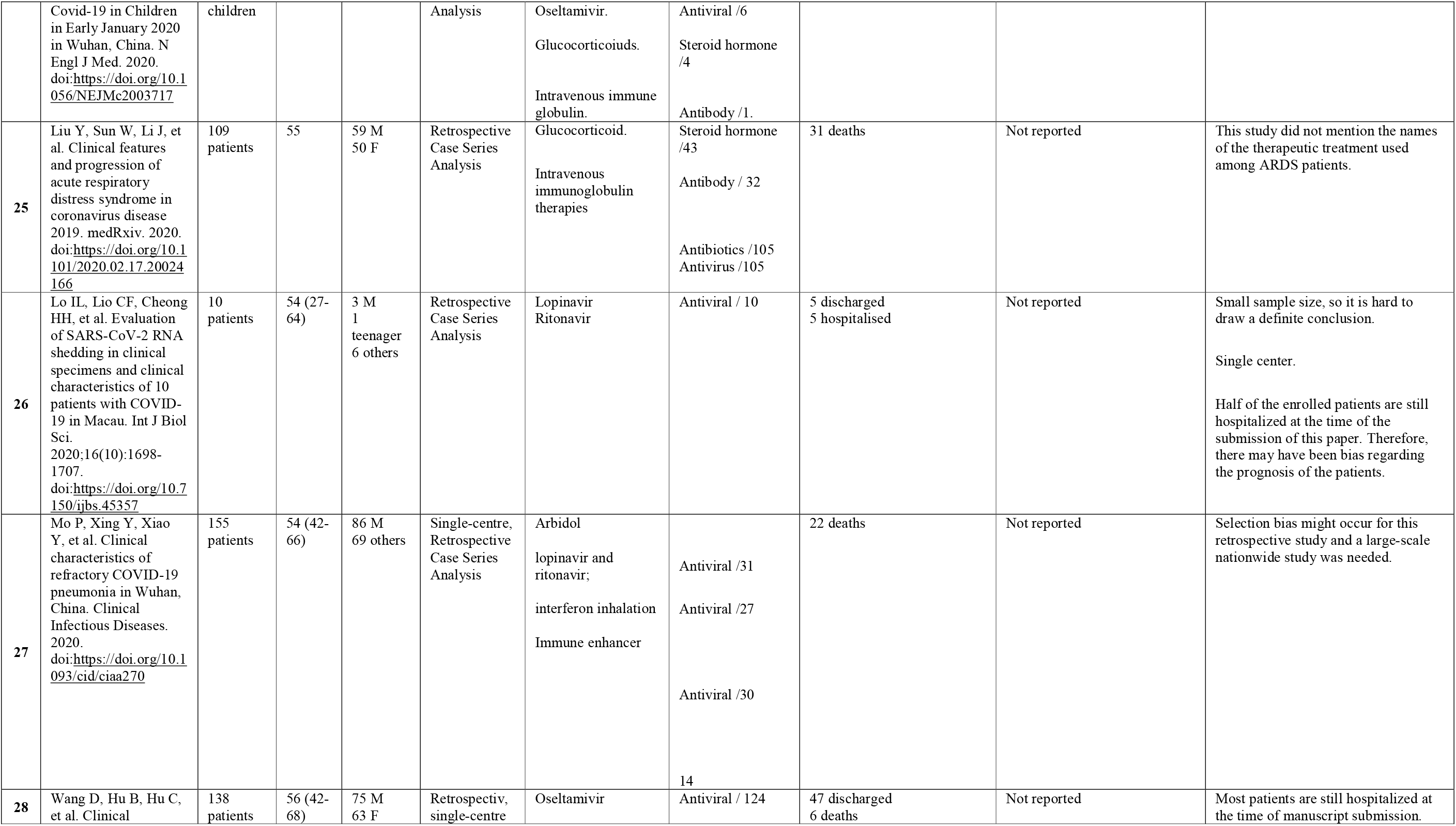
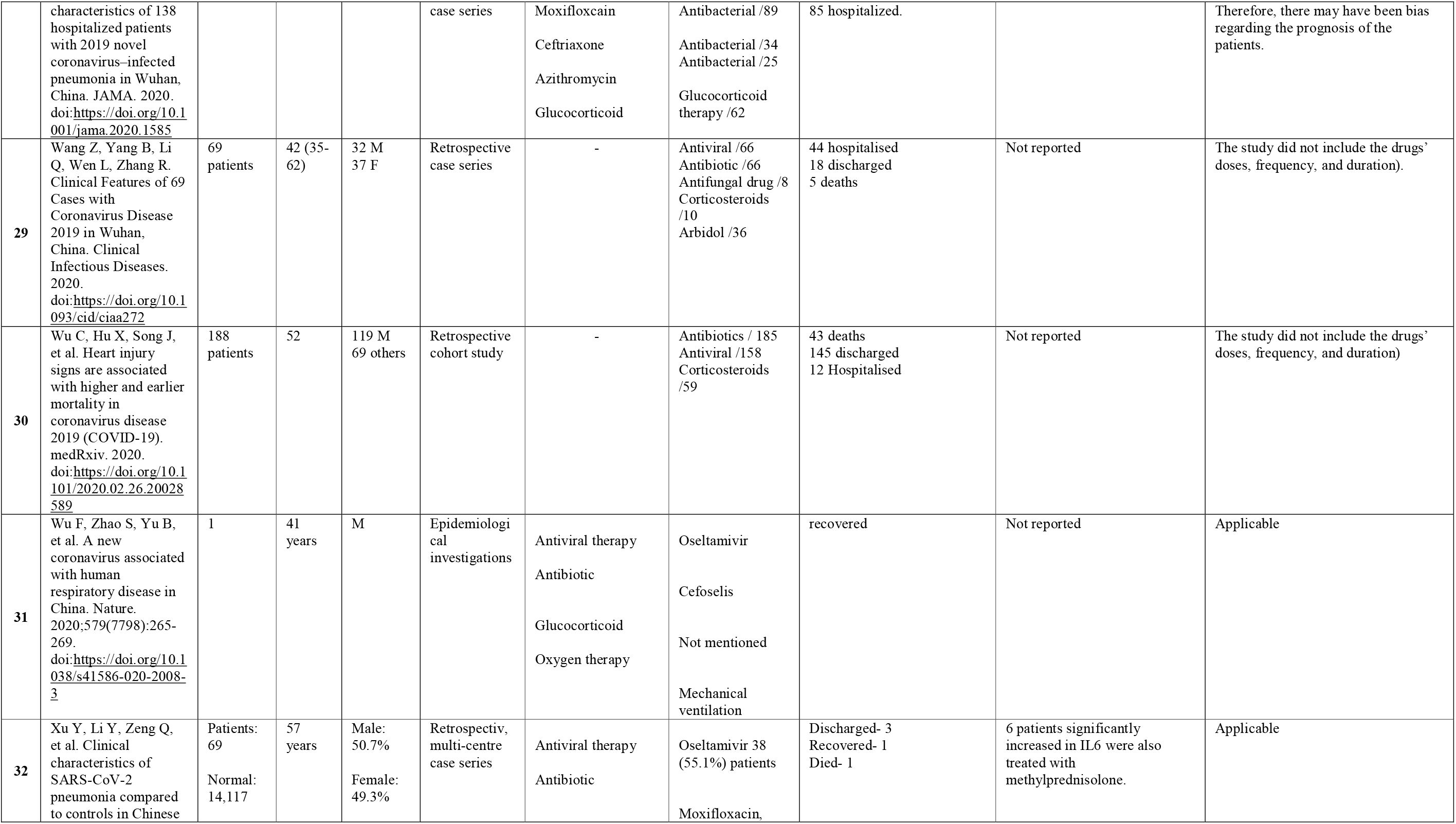
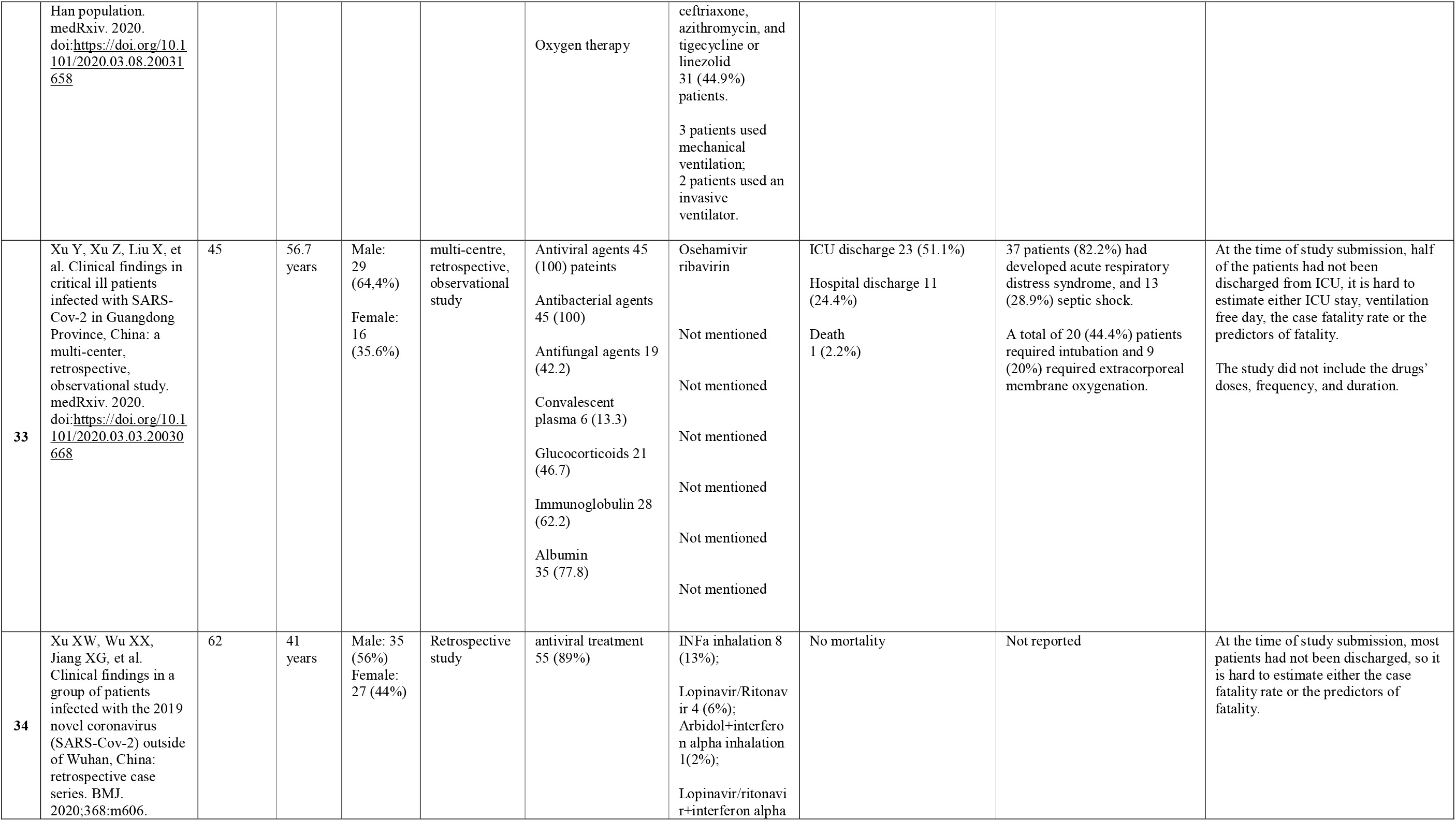
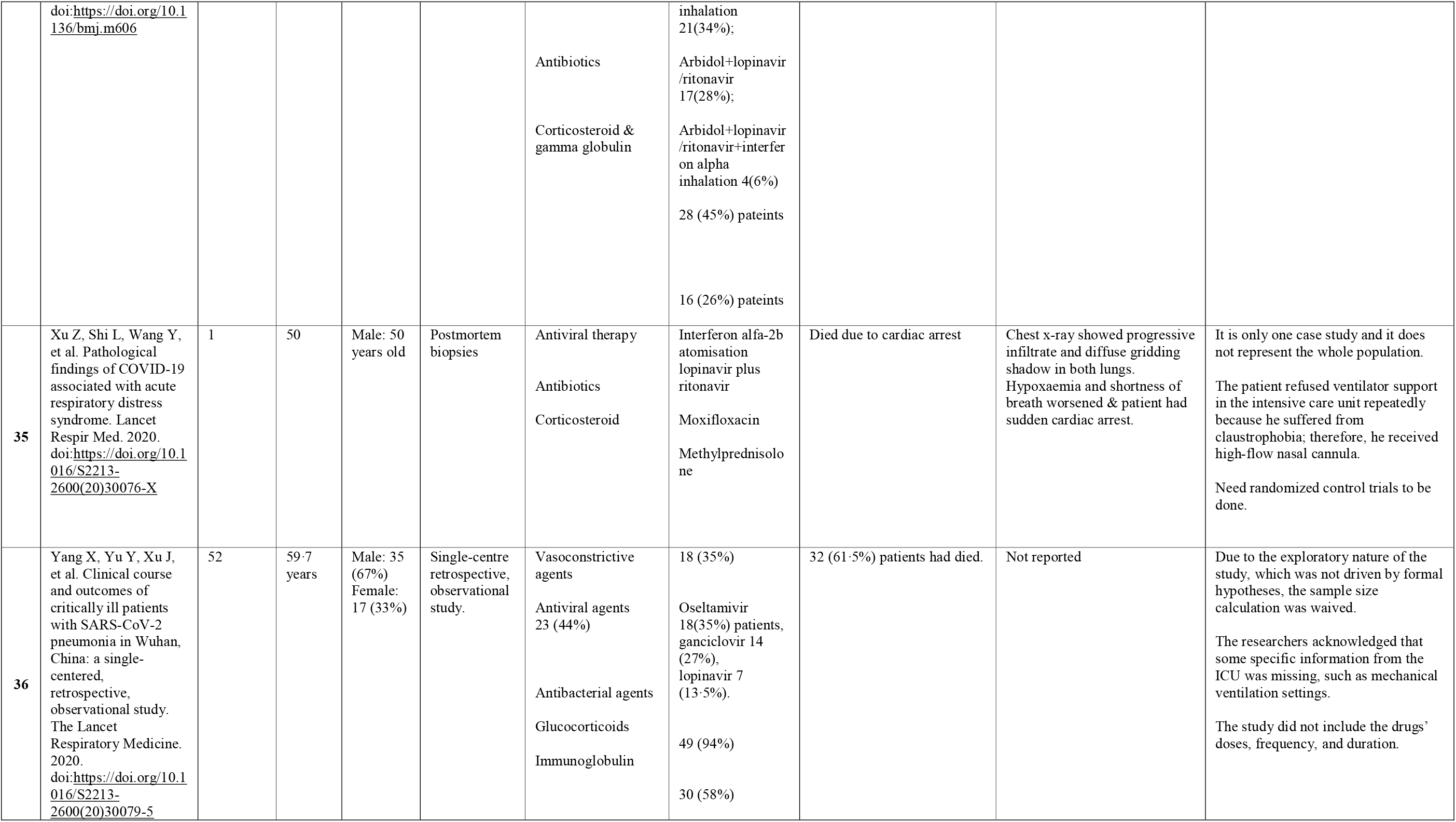
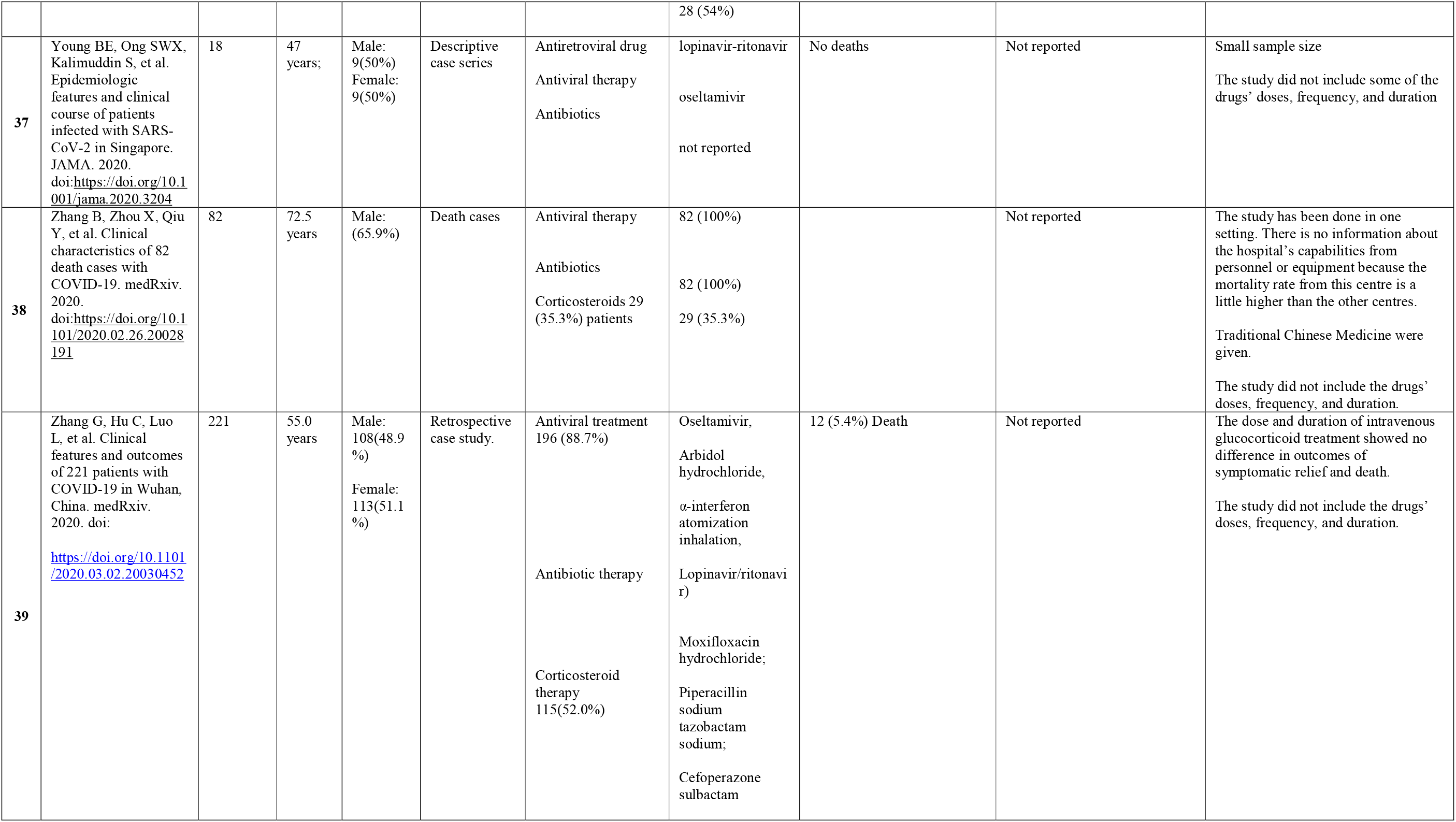
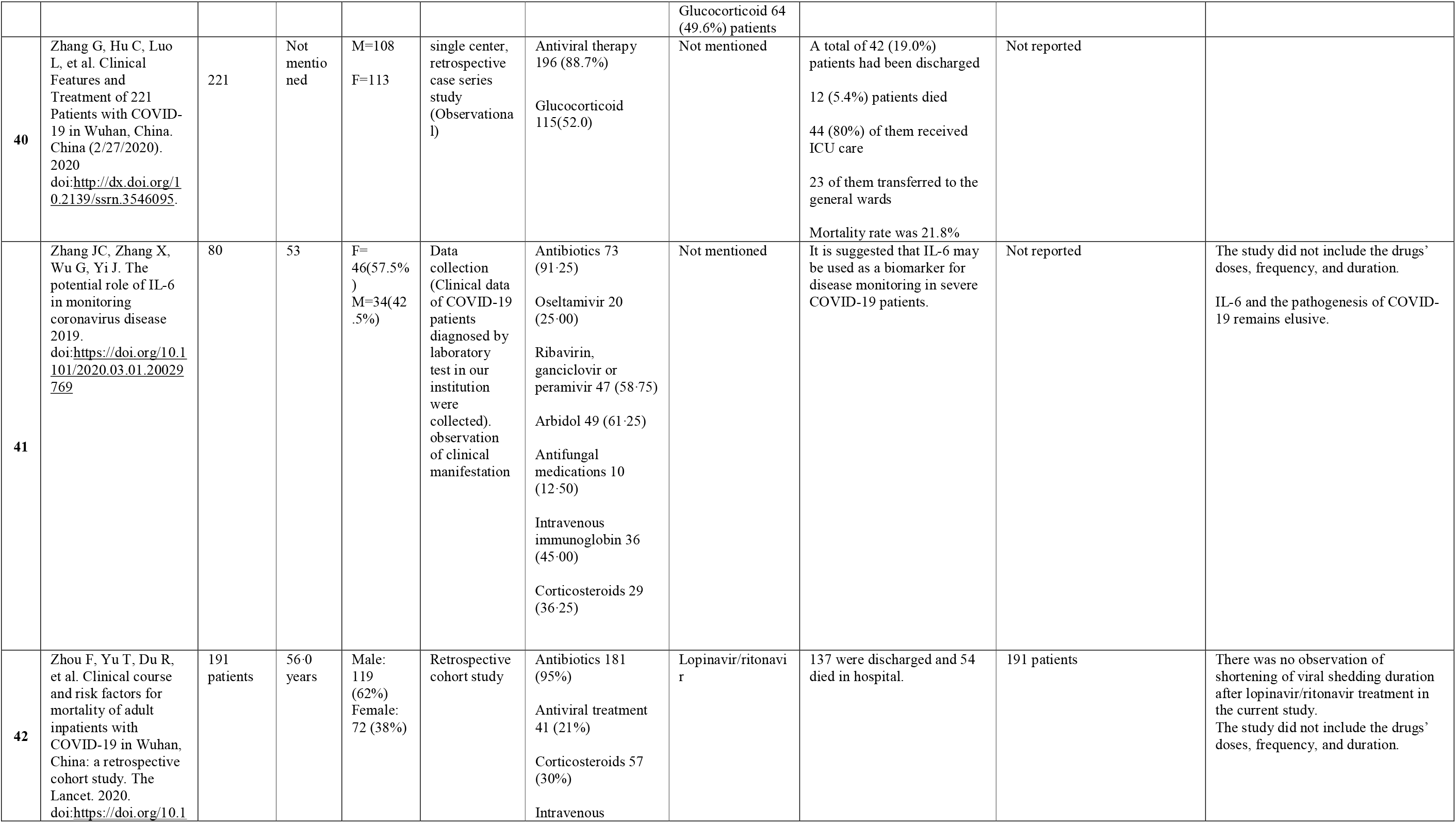
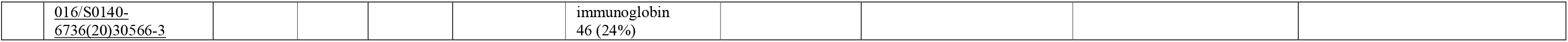
Data extraction from included papers

Forty-one studies were included, of which clinical trials (n=3), (case reports n=7), case series (n=10), retrospective (n=11) and prospective (n=10) observational studies.

Thirty-six studies were conducted in China (88%), Korea (n=1), USA n=1, France n=1, Singapore n=1, Macau n=1.

### Patients characteristics

Total Number of patients reported in this study were 8806: males (n=2111, 24%), females (n=3638, 41.3), other (n=2995, 34%), unspecified (n=62, 0.3%).

The mean of age was 50.8 years in 39 studies; the age was not specified in three studies.

### Reported therapeutics

The most common reported medicine in this systematic review was the anti-inflammatory medication corticosteroid (n=25) with different names and product characteristics, (corticosteroid n=21, methylprednisolone n=3, dexamethasone n=1), followed by the antiviral HIV medication Lopinavir (n=21), as combination lopinavir / ritonavir (n=18), alone (n-3), followed by the oseltamivir (n=16) and arbidol hydrochloride (n=8).

In terms of antibacterial medicines, moxifloxacin (n=4) and tigecycline were the most reported.

Convalescent plasma therapy was reported in one multi center retrospective observational study in (n=6) patients.

### The outcome of the treatment

The outcome of the therapeutics given varies between patients’ discharge and recovery, ongoing hospitalization, and mortality. Available data concerning this issue is shown in Table 3.

## Discussion

This is the first up to date review related the therapeutics used in COVID-patients in a systematic manner. As of March, twenty-sixth, 2020 and since the emergence of COVID-19, only forty-one research articles on COVID-19 and therapeutics were found eligible to be included in the current systematic review^(2,5,12-49),^ of which only three were clinical trials; most were either case reports, case series or prospective and retrospective observational studies. Systematic corticosteroid of different names and formulation was the most commonly mentioned, and used medication (n=25), followed by the antivirals Lopinavir (n=21), oseltamivir (n=16) and arbidol hydrochloride (n=8). The Convalescent plasma therapy was mentioned in one multi-center retrospective observational study and was administered to six patients.

Although quality assessment was applied to the included research articles, there was insufficient evidence from the articles identified in this review to conduct a meta-analysis. Nor was a subgroup analysis (adults and children, different formulations, dosages and duration) appropriate.

Most reported articles in this review are low quality, the design and the outcome of the studies are either incomplete or inconsistent, hence difficult to interpret the therapeutics in terms of efficacy and safety.

Despite these limitations, this is the first systematic review linked the therapeutics used in COVID-19 patients. Furthermore, the review provided up-to-date insight on the current therapeutics’ guidelines for the management of COVID-19 patients, most of reported medicines in this review were already in place in the USA, Saudi Arabia, Europe, and Egypt (Table1).

**Corticosteroids** were the first most commonly mentioned and used medicine in this review; however, it was deemed not recommended in any of the mentioned guidelines. The World Health Organization (WHO) and the United States Centers for Disease Control and Prevention (CDC), in the absence of conclusive scientific evidence, recommended that corticosteroids should not be routinely used in patients with COVID-19 for treatment of viral pneumonia or acute respiratory distress syndrome (ARDS) unless indicated for other conditions, such as asthma or chronic obstructive pulmonary disease (COPD) exacerbation, and septic shock^(5,50-51)^. Careful use of corticosteroids with low-to-moderate doses and short courses of treatment is advised. Hyperglycemia, hypernatremia, and hypokalemia are the most common adverse effects associated with corticosteroids’ use and should be routinely monitored ^(5,51)^.

**Lopinavir/ritonavir** is available as the brand name (Kaletra) and was the second most reported medicine in this review. Cao B and colleagues showed in their RCT negative outcomes of this HIV treatment for COVID-19 patients (Table 2) ^(30,52-54),^ no benefit was observed with lopinavir–ritonavir treatment beyond standard care in this study, nineteen patients from whom received the intervention died. However, some limitations were observed in the study, including the lack of blindness. RCT NCT04252885 and SOLIDARITY trial are ongoing to determine the efficacy in Lopinavir/ritonavir COVID-19 patients ^(52)^.

**Oseltamivir** was the third most mentioned medicine in this review, and sold under the brand name Tamiflu, it is used to treat influenza A and influenza B. Oseltamivir was recommended by WHO for people at high risk of infection for prevention of pandemic influenza. Guan W and colleagues in their retrospective observational study reported the use of oseltamivir in 1099 patients; however, the study was not able to provide any solid data on the effectiveness of oseltamivir in the prevention or treatment of COVID-19 patients. Study limitations were incomplete documentation of patients’ data and recall bias ^(55-56)^.

**Abidol hydrochloride** was the fourth most mentioned medicine in this review; it is a broad-spectrum inhibitor of influenza A and B virus, parainfluenza virus, and other types of viruses including hepatitis C virus. It is used in Russia and China, yet not approved for use in other countries ^(52)^. However, no conclusive evidence of its efficacy for COVID-19 was reported. In this review, it was reported together with favipiravir, which was approved recently for treatment of novel influenza on February 15, 2020 in China^(52)^.

**Chloroquine phosphate** and **Hydroxychloroquine** were reported in this review and showed favorable outcomes on the recovery of COVID-19 patients ^(6-7,57-60)^. The mechanism of action on viruses for these two medicines is probably the same effect. Chloroquine has been used for a long time to treat malaria and showed positive outcomes on patients. Furthermore, Hydroxychloroquine showed a significant effectiveness to kill intracellar pathogens such as Coxiella burnetii, the agent of Q fever ^(22)^. The French open label non-randomized clinical trial was promising and the first clinical trial of these medication on COVID-19 patients. The effect of Hydroxychloroquine was significant because it showed reduction in the viral load when it compared with the control group ^(22)^. Moreover, the effect of Hydroxychloroquine was significantly more effective when azithromycin was added to the patients according to their clinical needs. However, clinical follow-up and occurrence of adverse effects were not discussed in the paper; further work should be done on these medicines to reduce the spread of COVID-19 ^(57-59)^. Although these two medicines have shown promising activity against SARSCoV-2, there is a risk of arrhythmia associated with their requiring caution for use at higher cumulative dosages. Therefore, it is recommended that their use in suspected/confirmed COVID-19 is to be restricted to hospitalized patients. On March 30, 2020, has issued an emergency use authorization for chloroquine and hydroxychloroquine to treat patients hospitalized with Covid-19 ^(60)^.

**Convalescent plasma treatment** was mention once in this review, in a multi-center cohort research of 45 critically ill COVID-19 patients admitted to ICU in Wuhan. The findings showed that convalescent plasma was administered to six patients and no transfusion reactions occurred; however, the study could not provide adequate information about the efficacy of convalescent plasma, due to limited sample sizes and lack of randomized controlled group ^(61-62)^.

In fact, convalescent plasma therapy could be a promising method of treatment for COVID-19 patients, in a very recent study conducted in China (case series), showed that five critically ill patients with laboratory confirmed COVID-19 who had acute respiratory distress syndrome; their body temperature was normalized within 3 days in 4 of 5 patients after receiving plasma transfusion of recovered COVID-19 patients, their viral loads also decreased and became negative within 12 days after the transfusion; 3 out of 5 patients, were discharged from the hospital and were in stable condition at 37 days after transfusion ^(63)^.

On March 24, 2020 the US FDA has approved convalescent plasma treatment for investigational use under the traditional Investigational New Drug Applications (IND) regulatory pathway, and for **e**ligible patients who have confirmed COVID-19 and severe or immediately life-threatening conditions such as respiratory failure, septic shock, and/or multiple organ dysfunction or failure ^(64-65)^.

Notably there are some potential risks and ethical issues associated with their used, including increased thrombotic event risk that occur at the same administration date (0.04 to 14.9%), the lack of high quality research in this particular area, and the selectivity of donors with high neutralizing antibody titers ^(65)^.

## Conclusions

This is the first systematic review up to date, related to the therapeutics used in COVID-19 patients. Only 41 research articles on COVID-19 and therapeutics were found eligible to be included, most conducted in China, of which only three were clinical trials.

The anti-inflammatory medication corticosteroid was found to be the most mentioned and used medicine in these studies, despite the safety alert made by WHO and CDC, followed by antivirals medication Lopinavir, oseltamivir and arbidol hydrochloride.

Although further research is warranted as the amount of the evidence increases, but with the limited available data, this study presents the current picture of treatment modalities used for COVID-19. Propelling developments in treatment of COVID-19 are needed to be characterized in future studies.

## Data Availability

The data that support the findings of this study are available from the corresponding author, [MT],upon reasonable request.

